# *APOE4* genotype and *MAPT* haplotype modify repetitive head impact biomarkers in retired professional fighters

**DOI:** 10.1101/2025.09.19.25335803

**Authors:** Xiaowei Zhuang, Dietmar Cordes, Lynn Bekris, Edwin C. Oh, Charles Bernick

## Abstract

Long-term vulnerability to brain injury after repetitive head impacts (RHI) is poorly predicted *in vivo.* The wide heterogeneity of outcomes suggests that common genetic variants may act as modifiers of RHI-related neurodegeneration. Here, we tested whether *APOE* ε4 and *MAPT* H1/H2 haplotypes function as genetic modifiers of biomarker trajectories in 111 retired professional fighters from the Professional Athletes Brain Health Study (PABHS), the largest systematically characterized living cohort of fighters. RHI exposure was indexed by lifetime number of professional bouts. Primary outcomes included plasma GFAP (astroglial activation), NfL (axonal injury), and hippocampal volumes, measured cross-sectionally and longitudinally. Pre-specified linear models tested exposure × genotype interactions with false-discovery-rate correction. Across genotypes, greater fight exposure was associated with higher GFAP and smaller hippocampal volume. *APOE* ε4 modified longitudinal GFAP responses, with carriers showing stronger exposure-related increases than noncarriers, consistent with heightened astroglial reactivity. Conversely, the *MAPT* H2 haplotype mitigated associations between exposure, NfL, and hippocampal atrophy, suggesting partial protection against neuroaxonal injury and structural decline. Exploratory analyses showed *MAPT* H1/H1 enrichment among fighters meeting traumatic encephalopathy syndrome (TES) criteria (OR=3.33). Plasma p-tau231 was unrelated to exposure, indicating these effects are unlikely to reflect concurrent Alzheimer-type tau pathology. Together, these findings provide *in vivo* evidence that *APOE* and *MAPT* act as genetic modifiers of RHI-related brain injury, complementing postmortem evidence linking *MAPT* to chronic traumatic encephalopathy and highlighting the potential of genotype-informed monitoring in contact-sport populations.

**Significance Statement:** Repetitive head impacts (RHI) can lead to lasting brain damage, but not everyone exposed to RHI experiences the same outcomes. In retired fighters, we show that common genetic variants influence whether individuals are more vulnerable or resilient to brain inflammation and injury after repeated trauma. Specifically, APOE4 increased susceptibility, while a MAPT H2 variant appeared protective. These results provide the first in vivo evidence that inherited genetic differences modify long-term brain responses to RHIs. Incorporating genetic risk into studies of brain injury could enable earlier identification of at-risk individuals and support more personalized strategies for monitoring, prevention, and intervention.

## Introduction

Professional fighters are routinely exposed to repetitive head impacts (RHI) during both sanctioned matches and intensive training, substantially elevating their risk for long-term neurological conditions including chronic traumatic encephalopathy (CTE)^1,2^. RHI is a well-established primary driver of CTE pathology^3^, with greater exposure linked to altered blood-and brain-based biomarkers in professional fighters^4–6^. However, not all fighters with comparable RHI exposure develop the same degree of neurological impairment, suggesting that additional factors, such as genetic predisposition, may act as modifiers of susceptibility^2,7^. Defining genetic influences on RHI-related biomarkers could improve risk stratification, inform training practices, and guide development of personalized strategies to mitigate long-term neurological damage.

Several genes have been studied in this context^8^. The *Apolipoprotein E (APOE)* E4 allele, a well-established risk factor for sporadic Alzheimer’s disease (AD)^9,10^, has been linked to pathological mechanisms including neuroaxonal injury, neuroinflammation, neurometabolic changes and neurotoxicity, all of which are implicated in both single episode and repetitive head trauma^11,12^. Post-mortem analysis of 364 CTE brain donors demonstrated that *APOE4* was associated with more advanced CTE stages and increased phosphorylated tau (ptau) burden in individuals over 65, suggesting a potential role of *APOE4* in CTE pathogenesis^7^. Yet its contribution to other RHI-related pathologies, such as chronic neuroinflammation or axonal injury, remains unclear. In vivo studies of APOE4 in fighters and athletes have produced inconclusive results^13–15^, often limited by small samples, cross-sectional designs, and the inclusion of both active and retired athletes, which reduces statistical power and generalizability. Beyond *APOE*, the *microtubule-associated protein tau (MAPT)* gene is also of interest.

*MAPT* encodes the tau protein that becomes hyperphosphorylated and forms neurofibrillary tangles (NFTs) in various neurodegenerative disorders including CTE^16^. The *MAPT* H2 haplotype has been reported to confer protection against certain sporadic and primary tauopathies like progressive supranuclear palsy (PSP), compared to the more common H1 haplotype^16–18^. Although tauopathy is central to CTE, limited studies have examined the influence of *MAPT* haplotype on CTE neuropathology or RHI-related biomarkers. One study found a trend towards a higher frequency of *MAPT* H1/H1 carriers in athletes with CTE compared to those without^19^. More recently, a structural haplotype in the 17q21.31 region (encompassing *MAPT*) was significantly associated with dementia and post-mortem tau-burdens in CTE-affected brain regions^20^. Nevertheless, to our knowledge, no studies to date have investigated the influence of *MAPT* haplotype on *in vivo* biomarkers reflecting various RHI-related neuropathological processes.

The Professional Athletes Brain Health Study (PABHS), a longitudinal study of RHI effects in living fighters^21^, provides a unique opportunity to fill this gap. Prior work using PABHS data has linked fighting exposures to increased plasma levels of neurofilament light chain (NfL) and glial fibrillary astrocytic protein (GFAP) levels^22–25^, as well as reduced hippocampal volumes^4,5,25,26^. GFAP reflects astrocytic activation and neuroinflammation^27^, NfL is a marker of axonal injury^28^, and hippocampal volume loss indicates neuronal degeneration. Together, these biomarkers capture processes central to RHI-related neurodegeneration. With the recent addition of genotyping data, PABHS now enables direct investigation of whether genetic factors moderate these biomarker changes. In this study, we tested whether *APOE4* and *MAPT* haplotype act as genetic modifiers of these biomarker trajectories. We hypothesized that greater fight exposure would associate with adverse biomarker changes post-retirement and that *APOE4* and *MAPT* H1/H1 carriers would show greater vulnerability compared to their counterparts.

## Methods Participants

This study is part of the PABHS approved by the Cleveland Clinic Institutional Review Board (IRB protocol #10-944). Details of PABHS have been previously reported^21,22^

We included fighters (boxers, mixed martial artists (MMA), and martial artists (MA)) with available genotyping data and at least one visit classified as post-retirement. We focused on retired fighters to rule out effects of acute head trauma on biomarker levels. Fighting histories, including numbers of professional fights (nof_pro) and last fighting dates, were verified using Boxrec (www.boxrec.com) for boxers and Sherdog (www.sherdog.com) for MMAs and MAs. Cumulative RHI exposure was quantified as the nof_pro throughout the career. We defined *qualified* visits as occurring at least two years post-retirement. The final analytical sample comprised 111 retired fighters with blood biomarkers and MRI data collected at *qualified* visits. (**blue boxes in** **Fig. 1**).

**Figure 1.**
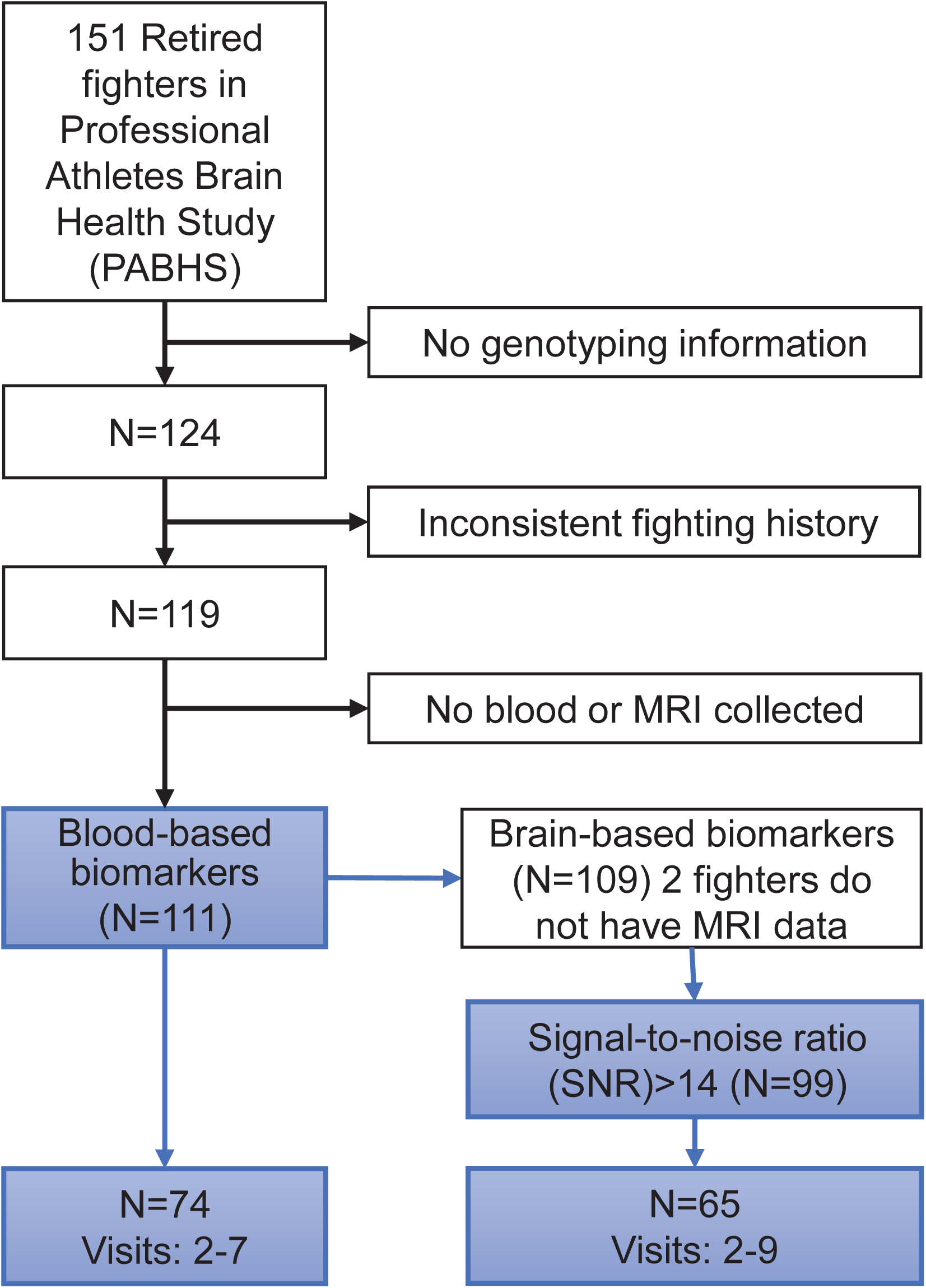
Flow diagram of participant inclusion. Of 151 retired fighters enrolled in the Professional Athletes Brain Health Study (PABHS), exclusions were due to missing genotyping, inconsistent fight histories, or absence of biomarker/MRI data. The final Core Baseline Cohort comprised 111 fighters with blood or MRI biomarkers, and a Core Longitudinal Cohort of 74 fighters contributed ≥2 qualified post-retirement visits. MRI scans were required to pass a signal-to-noise ratio (SNR) >14. Abbreviations: MRI, magnetic resonance imaging; SNR, signal-to-noise ratio; PABHS, Professional Athletes Brain Health Study.

### Real-time polymerase chain reaction (PCR) for *APOE* and *MAPT* genotyping

*APOE* and *MAPT* single nucleotide polymorphisms (SNPs) were genotyped using allelic discrimination PCR in the Bekris Lab at the Cleveland Clinic main campus^29^. *APOE* genotype was determined by SNPs rs429358 and rs7412, while *MAPT* haplotypes were determined via SNP rs1800547 that tags to haplotype^30^. Retired fighters were categorized into *APOE4* carriers (i.e. C/C or C/T at rs429358) or non-carriers (i.e. T/T at rs429358), and *MAPT* H1/H1 carriers (A/A at rs1800547) or H2 carriers (A/G or G/G at rs1800547), respectively.

### Fighting-related blood- and brain-based biomarkers

Blood-based biomarkers: Venipuncture was performed at fighters’ annual visit. Plasma NfL and GFAP concentrations were measured using ultrasensitive molecule array assays (Quanterix, Billerica, MA, USA) at the Clinical Neurochemistry Laboratory, Sahlgrenska University Hospital (Mölndal, Sweden). A subset of fighters had plasma ptau-231 measured following published protocols^31^. All procedures were performed by board-certified laboratory technicians blinded to clinical data and followed manufacturer specifications.

Brain-based biomarkers: High-resolution T1-weighted structural MRI was acquired on a 3T Siemens scanner during fighters’ annual visit using a standard 3D MPRAGE sequence (TR=2300ms, TE=2.98ms, TI=900ms, flip angle=9°, in-plane resolution=1mmx1mm, slice thickness=1.2mm). Hippocampal volumes were calculated using the FreeSurfer 6.0 longitudinal processing pipeline^32^. A signal-to-noise ratio (SNR) was computed for every scan using the FreeSurfer quality assessment tool, and only data with a SNR>14 was utilized in the following analyses (**Fig. 1**).

### Statistical analysis

Demographic differences between genetic groups were assessed using Chi-square tests for categorical variables and two-sample t-tests for continuous variables. Linear regression models were utilized to evaluate the genetic effect on the relationship between nof_pro and each biomarker. For cross-sectional analysis, biomarker measure from the first *qualified* visit of each retired fighter (i.e., Baseline) served as the dependent variable; while nof_pro, genotype, and their interactions were the independent variables. Additional covariates included sex, age at measurement, years of education, race and types of fighters. Total intracranial volume was included as a covariate brain regional volume analysis. For longitudinal analysis, the annual rate of change for each biomarker was first estimated for retired fighters with multiple *qualified* visits. The same linear regression model was applied to evaluate the genetic effect on biomarkers’ annual rate of change.

For measures showing an interaction effect between genotype and nof_pro (i.e., p-value for interaction term ≤0.05), post-hoc partial correlations (r) between nof_pro and the biomarker or biomarker annual changing rate were calculated within each genotype group, adjusting for covariates using regression coefficients estimated in the regression model. In addition, false discovery rate method (FDR-p) was utilized to correct for total 48 multiple corrections, given that we were interested in effects of fighting exposure, genotype and interaction terms on both cross-sectional and longitudinal blood- and brain-based biomarkers (i.e., 3x2x2x4).

### Results Participants

The Core Baseline Cohort included 111 retired fighters (91.9% male; mean [SD] age, 46.2 [9.5]) with verified fight histories, genotyping, and blood or MRI data passing quality control (**Table 1**). Race differed significantly between *MAPT* groups, with H2 carriers rare among African American participants (p=0.002). *MAPT* H1/H1 carriers were more likely to meet consensus criteria for traumatic encephalopathy syndrome (TES; OR=3.33, p=0.007). A Core Longitudinal Cohort of 74 fighters (mean [SD] follow-up 5.2 [2.1] years) contributed biomarker trajectories (**Supplementary Table 1**). Twelve participants (2 with no MRI, 10 failing SNR QC) were excluded from hippocampal analyses.

**Table 1.** Characteristics of 111 retired fighters with genotyping and biomarker data, stratified by *APOE4* carrier status and *MAPT* haplotype. Values are reported as mean [SD] unless otherwise specified. TES = traumatic encephalopathy syndrome. Group comparisons were tested using t-tests for continuous variables and χ² tests for categorical variables.

| *APOE* | E4-non carriers<br>(mean [std]) | E4 carriers<br>(mean [std]) | Between<br>group<br>differences<br>(p-value) |
| --- | --- | --- | --- |
| Number of retired fighters<br>(N=111) | 87 | 24 |  |
| with ≥2 qualified visits<br>(N=74) | 57 | 17 |  |
| Sex | 7 Female<br>80 Male | 2 Female<br>22 Male |  |
| age@1<sup>st</sup> available<br>measure (Baseline) | 46.47 [9.41] | 45.08 [10.04] |  |
| Race | 60 White<br>21 African American<br>6 Other | 13 White<br>7 African American<br>4 Other |  |
| Years of education | 13.20 [2.16] | 13.29 [2.26] |  |
| Number of professional<br>fights throughout career | 31.38 [17.64] | 24.54 [14.81] |  |
| Types of fighters | 62 Boxers<br>1 Martial Artist<br>21 Mixed Marial Artists<br>3 Mixed fighters | 17 Boxers<br>6 Mixed Marial Artists<br>1 Mixed fighter |  |
| Retired Fighters type | 66 enrolled as retired fighters<br>21 transitioned | 16 enrolled as retired fighters<br>8 transitioned |  |
| TES status till latest visit | 45 TES | 10 TES |  |
| NfL (pg/ml) | 8.46 [4.41]<br>Range [3.45, 31.96] | 8.36 [6.32]<br>Range [3.99,36.07] |  |
| GFAP (pg/ml) | 100.05 [56.90]<br>Range [33.60,421.96] | 95.16 [54.48]<br>Range [29.10, 244.00] |  |

Table 1
| *MAPT* | H1/H1 carriers<br>- A/A at rs1800547<br>(mean [std]) | H2 carriers<br>- A/G or G/G<br>at rs1800547<br>(mean [std]) | Between<br>group<br>differences<br>(p-value) |
| --- | --- | --- | --- |
| Number of retired fighters<br>(N=111) | 80 | 31 |  |
| with ≥2 qualified visits<br>(N=74) | 54 | 20 |  |
| Sex | 6 Female<br>74 Male | 3 Female<br>28 Male |  |
| age@1<sup>st</sup> available<br>measure (Baseline) | 46.06 [9.66] | 46.45 [9.30] |  |
| Race | 45 White<br>27 African American<br>8 Other | 28 White<br>1 African American<br>2 Other | 0.002 |
| Years of education | 13.11 [2.19] | 13.52 [2.13] |  |
| Number of professional<br>fights throughout career | 30.58 [16.91] | 28.16 [18.21] |  |
| Types of fighters | 57 Boxers<br>1 Martial Artist<br>21 Mixed Martial Artists<br>1 Mixed fighter | 22 Boxer<br>6 Mixed Martial Artist<br>3 Mixed fighters |  |
| Retired Fighters type | 58 enrolled as retired<br>fighters<br>22 transitioned | 24 enrolled as retired<br>fighters<br>7 transitioned |  |
| TES status till latest visit | 46 TES | 9 TES | 0.007 |
| NfL (pg/ml) | 8.34 [4.39]<br>Range [3.55,31.96] | 8.72 [6.01]<br>Range [3.45,36.07] |  |
| GFAP (pg/ml) | 98.31 [47.29]<br>Range [29.10, 232.00] | 100.74 [75.35]<br>Range [33.60,421.94] |  |

### Genetic influence on neuroinflammation biomarker profiles

Significant positive associations were observed between nof_pro and GFAP levels at the first *qualified* post-retirement visit (i.e., Baseline) in models with either *APOE* (β=1.324, SE=0.349, p=0.0003, FDR-p=0.012) or *MAPT* (β=1.021, SE=0.387, p=0.010, FDR-p=0.077, **Fig. 2**) as genetic factors. Higher number of fights was correlated with higher GFAP levels across all genetic subgroups (solid lines in last columns).

**Figure 2.**
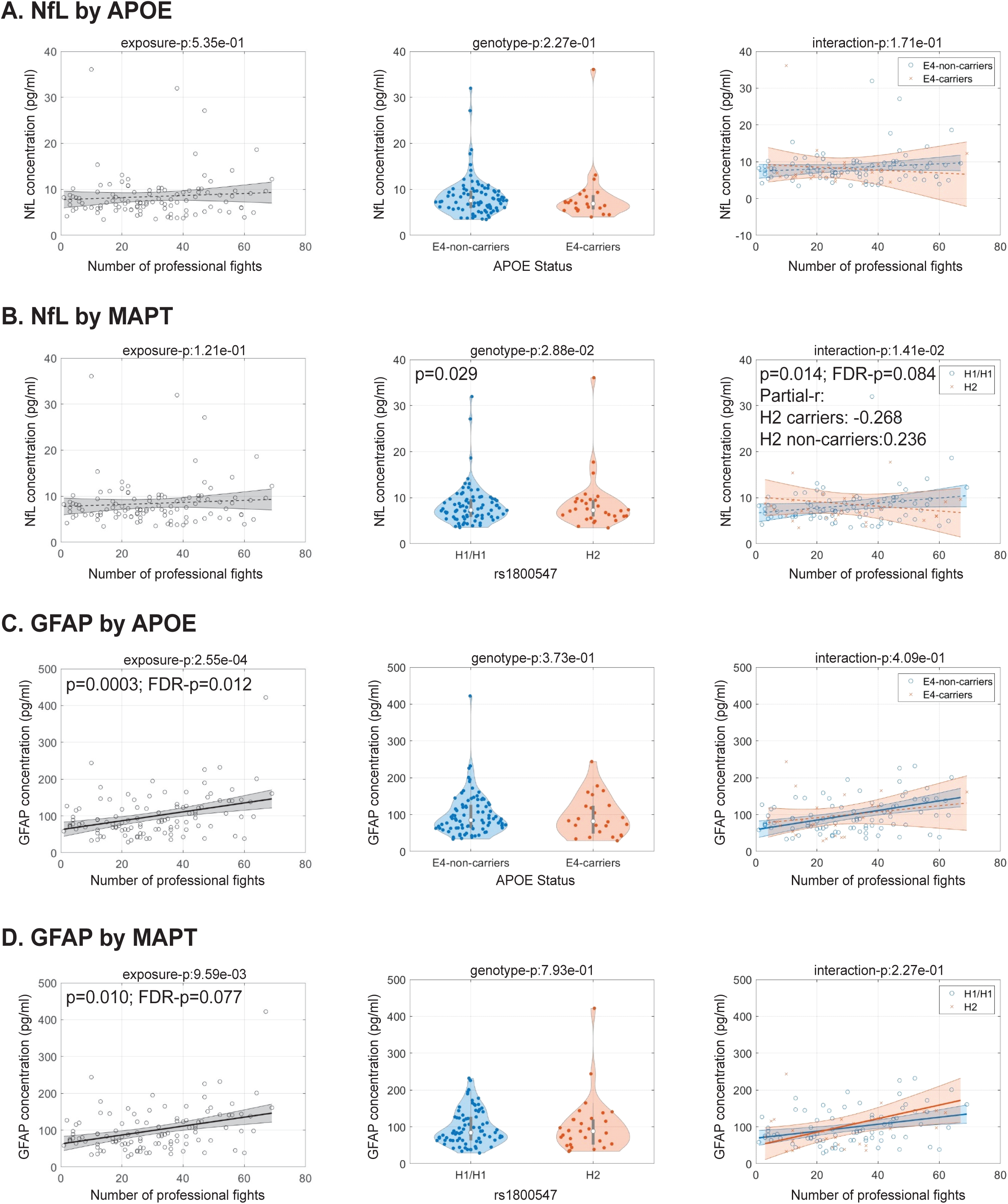
Cross-sectional associations of lifetime fight exposure (x-axis) with plasma NfL (top panels) and GFAP (bottom panels) at the first qualified post-retirement visit. Results are stratified by (A, C) *APOE4* status and (B, D) *MAPT* haplotype. Non-carriers (*APOE4*– or *MAPT* H1/H1) are shown in blue; carriers (*APOE4*+ or *MAPT* H2) in red. Regression lines reflect associations with 95% confidence interval shading. Violin plots show baseline biomarker distributions by genotype. For models with significant moderation (p<0.05), adjusted partial correlations (r) are reported. Solid regression lines indicate significant partial correlations in individual genotype groups. FDR correction was applied across cross-sectional and longitudinal analyses.

While longitudinal GFAP changes did not significantly correlate with fighting exposure across the entire cohort, *APOE4* carrier status significantly moderated this association (β=0.857, SE=0.296, p=0.005, FDR-p=0.049, **Fig. 3**). *APOE4* carriers showed a stronger positive relationship between nof_pro and longitudinal GFAP increases (partial-r=0.497) compared to non-carriers (partial-r=-0.133). Approximately 75.34% of retired fighters exhibited GFAP increases during the observation period.

**Figure 3.**
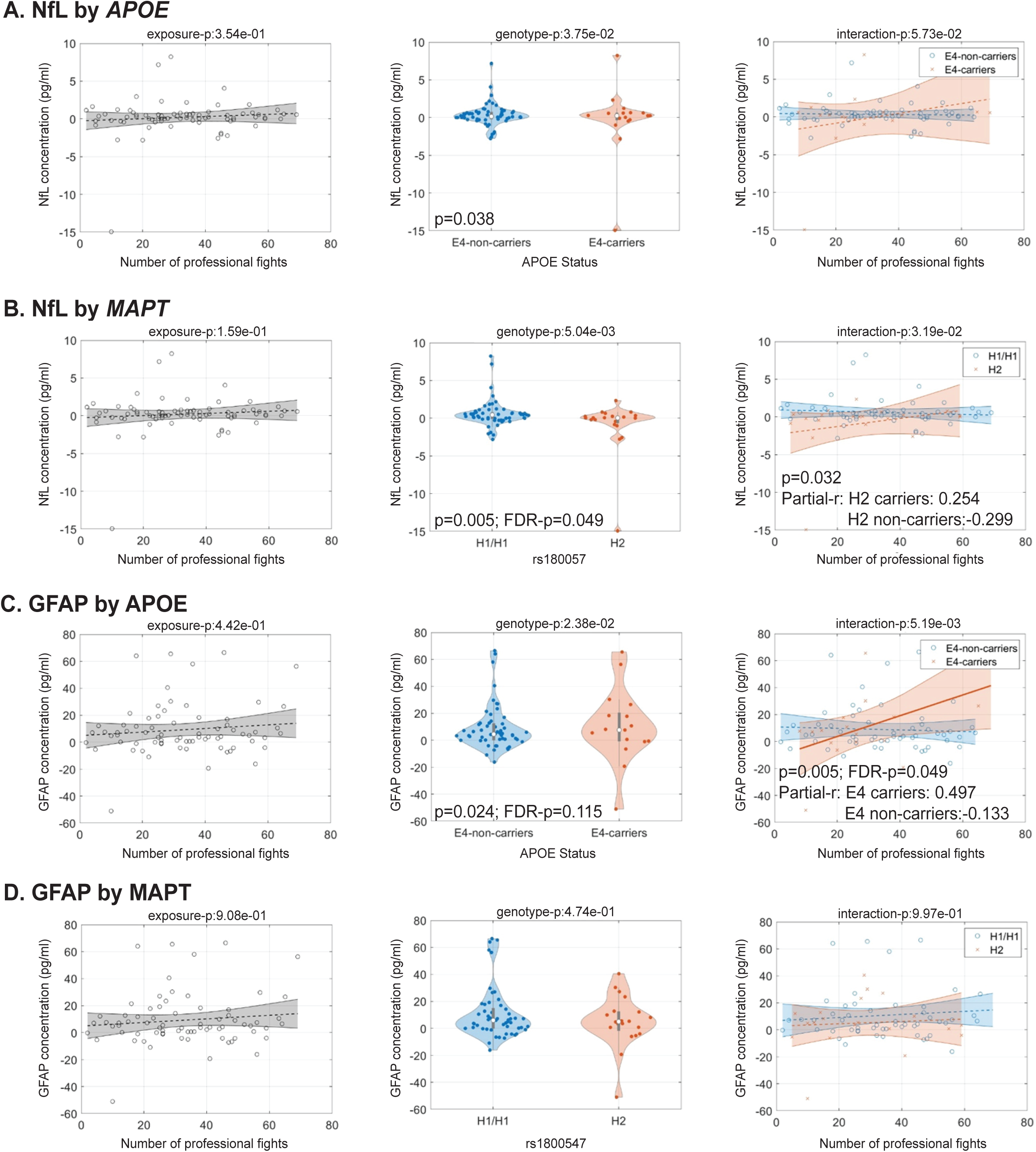
Longitudinal associations of lifetime fight exposure (x-axis) with annualized change in plasma NfL (top panels) and GFAP (bottom panels). **Results are stratified by (A,** C) *APOE4* status and (B, D) *MAPT* haplotype. Non-*carriers (APOE4– or MAPT H1/H1) are* shown in blue; carriers (*APOE4*+ or *MAPT* H2) in red. Regression lines reflect associations with 95% confidence interval shading. . Violin plots display average biomarker change per year by genotype group. Moderation effects with p<0.05 are annotated with partial correlations (r) adjusted for covariates. Solid regression lines indicate significant partial correlations in individual genotype groups. FDR correction was applied across families of analyses.

### Genetic influences on neuroaxonal injury biomarker profiles

Baseline NfL levels did not correlate with fighting exposure across the entire cohort. The *MAPT* H2 significantly moderated this relationship (β=-0.143, SE=0.057, p=0.014, FDR-p=0.084, **Fig. 2**), with *MAPT* H1/H1 carriers showing a positive association (partial-r=0.236), while H2 carriers exhibited a negative association (partial-r=-0.268). *MAPT* further moderated the relationship between nof_pro and longitudinal NfL changes at trend level significance (β=0.081, SE=0.037, p=0.032, FDR-p=0.118, **Fig. 3**). In addition, *MAPT* H2 carriers showed significantly attenuated longitudinal NfL increases compared to H1/H1 carriers (β=-3.543, SE=1.219, p=0.005, FDR-p=0.049). Around 40% of *MAPT* H2 carriers (8/20) showed decreasing NfL levels over time, with this NfL recovery effect diminishing as nof_pro increased.

Two retired fighters did not undergo MRI, and structural MRI scans from ten fighters at their first *qualified* post-retirement visit failed quality control. Therefore, these 12 fighters were not included in the analysis of hippocampal volumes. Nof_pro was significantly associated with reduced left hippocampal volumes (β=-15.424, SE=5.358, p=0.005, FDR-p=0.049), with this negative relationship consistent across all genetic subgroups (solid lines in last column plots in **Supplementary** Fig. 1). No significant genetic moderation effects were observed on cross-sectional hippocampal volumes.

Longitudinal analysis demonstrated that greater fighting exposure significantly associated with accelerated left hippocampal volume loss (β=-1.035, SE=0.319, p=0.002, FDR-p=0.048, **Fig. 4**). This relationship was further moderated by *MAPT* haplotype (β=1.250, SE=0.539, p=0.024, FDR-p=0.115), with H1/H1 carriers exhibiting more pronounced nof_pro-related volume loss compared to H2 carriers (partial-r=-0.496 vs. 0.105). Similar moderation patterns were observed for the right hippocampal volume loss (β=1.344, SE=0.625, p=0.044, **Fig. 4**).

**Figure 4.**
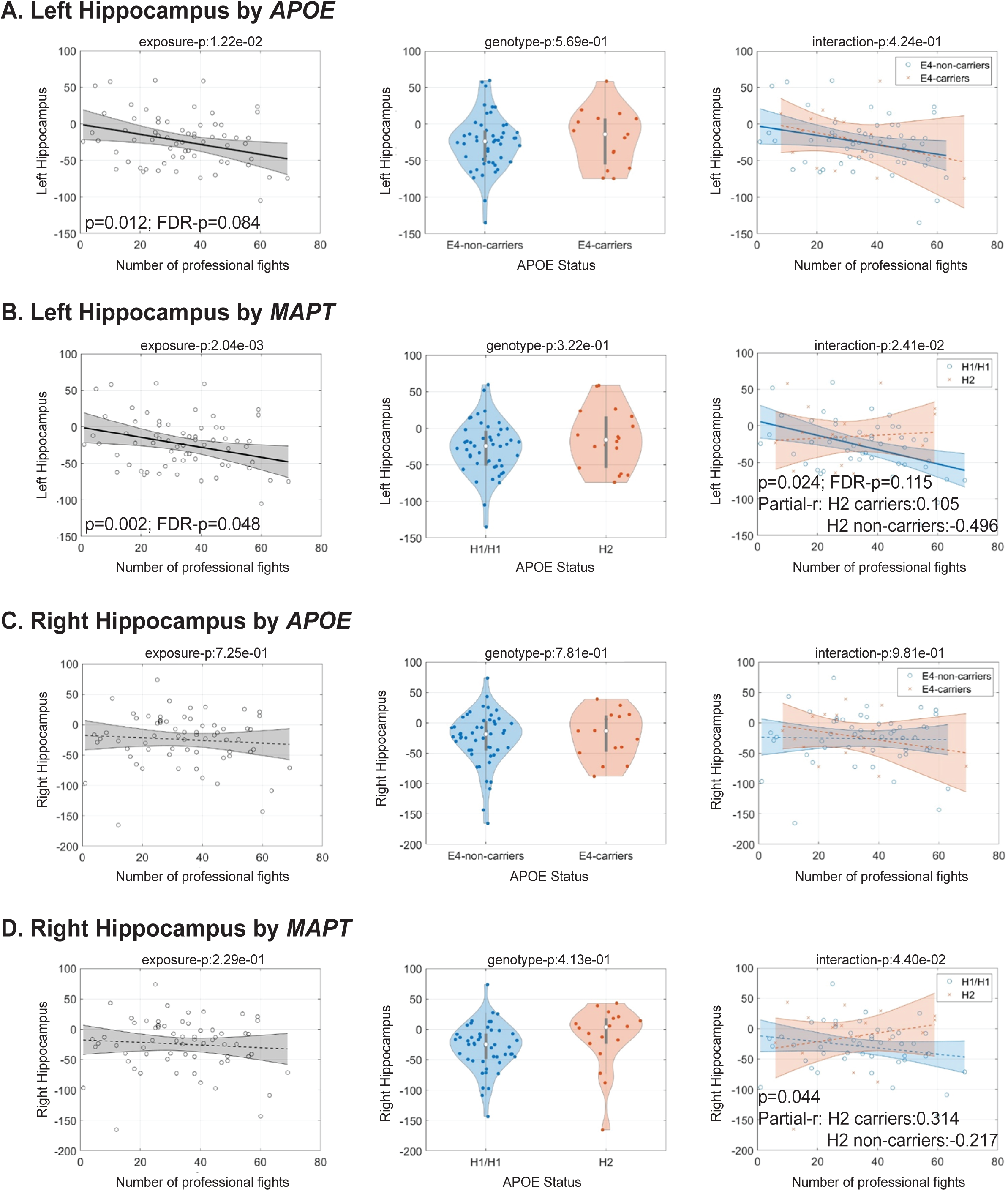
Longitudinal associations of lifetime fight exposure (x-axis) with annualized change in hippocampal volume (y-axis), stratified by *APOE4* status (A, C) and *MAPT* haplotype (B, D). Left hippocampus (top panels) and right hippocampus (bottom panels) are shown separately. Non-carriers *(APOE4– or MAPT H1/H1)* are plotted in blue; carriers (*APOE4*+ or *MAPT* H2) in red. Regression lines with 95% CI shading depict exposure-outcome relationships. Partial correlations are reported when interaction p<0.05. Solid regression lines indicate significant partial correlations in individual genotype groups. FDR correction was applied across cross-sectional and longitudinal analyses. .

## Discussion

In this study of 111 retired professional fighters, we examined whether the *APOE4* allele and *MAPT* H2 haplotype modify the relationship between repetitive head impact (RHI) exposure and *in vivo* biomarkers of neurodegeneration. Across the cohort, greater fighting exposure was associated with elevated baseline GFAP, reduced hippocampal volume, and accelerated hippocampal atrophy, reinforcing the link between RHI and both neuroinflammatory and neurodegenerative processes. Importantly, genetic variation moderated these associations: *MAPT* H2 carriers exhibited relatively preserved hippocampal volumes and attenuated NfL elevations, while *APOE4* carriers showed steeper longitudinal GFAP increases, suggesting heightened astroglial reactivity.

### Effect of RHI and *APOE4* on neuroinflammation

GFAP is a well-established biomarker of astrocytic activation and neuroinflammation^33^. Following severe TBI, GFAP levels would peak during a 12-hour window and may remain elevated into the chronic phase due to the persistent astrogliosis^34,35^. Meta-analyses suggest that *APOE4* carriers experience worse clinical and neuropsychological outcomes months to years after TBI, though with a small effect-size^11,36,37^, potentially through mechanisms including exacerbated direct neurotoxicity, modulation of tau pathology, disruption of the blood-brain barrier, and increased inflammation and oxidant injury^11^.

In the context of RHI, sustained GFAP elevation likely result from subsequent head traumas occurring before complete recovery to homeostasis from prior injuries, leading to prolonged astroglia activation and persistent neuroinflammation, which could be further exacerbated in *APOE4* carriers. Previous PABHS data demonstrated elevated GFAP levels in both active and retired fighters compared to controls^23^(**Supplementary** Fig. 3), corroborating these RHI-related neuroinflammatory changes. Our finding that a greater number of professional bouts associates with higher baseline GFAP levels approximately 10.8 years post-retirement, regardless of *APOE4* status, additionally highlight the dose-dependent and enduring nature of RHI-related neuroinflammatory responses. Importantly, *APOE4* status influenced the longitudinal trajectory of GFAP, with only *APOE4* carriers demonstrating a significant positive relationship between fight number and GFAP increases over time (**Fig. 3**). This finding supports our hypothesis that *APOE4* carriers may experience more pronounced neuroinflammatory responses to RHI compared to non-carriers.

However, both elevated GFAP and *APOE4* carrier status are associated with increased AD risks^10^. To address the possibility that underlying AD pathology might explain these findings, we conducted additional analyses in a subset of fighters with measured plasma ptau-231, a validated AD biomarker that correlates with post-mortem AD pathology and in vivo amyloid/tau positivity^31^. We found no association between fighting exposure and either baseline ptau-231 levels or longitudinal ptau-231 changes in either *APOE4* carriers or non-carriers (**Supplementary** Fig. 2), suggesting the observed GFAP findings are not driven by coexisting AD pathology in this cohort.

Collectively, these results indicate that *APOE4* may augment the neuroinflammatory response to RHI and suggest that elevated blood GFAP levels, when considered alongside *APOE4* genotype, may help identify retired fighters at increased risk for CTE-related neurodegeneration.

### Effect of RHI and *MAPT haplotype* on neuroaxonal damage

Blood-based NfL is an established biomarker of neuroaxonal injury^28^. Previous studies have documented acute NfL elevations following head trauma, such as after a single bout in Olympic boxers^38^, and throughout a season in American football players^39^. Our previous PABHS investigation demonstrated elevated NfL levels in active but not retired fighters^22,23^. Consistent with these reports, the present study found no significant associations between cumulative fighting exposure and either cross-sectional NfL levels or longitudinal NfL changes across the entire retired cohort. Instead, we observed that *MAPT* haplotype significantly moderates these associations (**Figs 2 and 3**), suggesting that while NfL effectively reflects acute neural injury during active RHI exposure, its utility in characterizing long-term neurobiological changes post-retirement may depend on genetic factors.

The *MAPT* H1 haplotype has been strongly associated with various primary tauopathies such as PSP ^16–18^. Sub-haplotypes or structural variants such as H1c and H1β1γ1 have been further linked to increased risks of AD^40^ and CTE^20^, respectively. In our cohort, a significantly greater proportion of retired fighters diagnosed with TES were *MAPT* H1/H1 carriers (odds ratio=3.33, *p*=0.007; **Table 1**), highlighting the potential role of *MAPT* in modulating susceptibility to neurodegeneration following RHI^19^. Our analysis provides novel *in vivo* evidence for a protective role of the H2 haplotype against long-term axonal degenerations, with H1/H1 carriers showing a positive association between fighting exposure and NfL levels, while H2 carriers demonstrated a negative association (**Fig 2**). This protective effect was further supported by the longitudinal NfL variations, where a substantial proportion of H2 carriers with lower fighting exposure exhibited decreasing NfL concentrations over time, potentially indicating neurobiological recovery post-retirement ^6^. Yet, this protective effect appeared to diminish with increasing fighting exposure, suggesting a potential RHI-dependent loss of resilience (**Fig. 3**).

We further examined this moderation effect using hippocampal volume as an anatomical marker of neurodegeneration, which is also known to be vulnerable in CTE^41^. While the hippocampus showed reduced volumes with greater fighting exposure across all genetic subgroups, longitudinal analyses revealed that only *MAPT* H1/H1 carriers exhibited significant hippocampal atrophy with increasing exposure (**Fig. 4**). These convergent findings suggest the *MAPT* H2 haplotype confers neuroprotective effects against RHI-related structural and molecular neurodegeneration, with important implications for identifying individuals at differential risk and developing targeted monitoring strategies.

### Limitations

We used the number of professional fights (nof_pro) as a proxy for cumulative RHI exposure in retired fighters. While previous research from PABHS has validated nof_pro as a reasonable proxy for fighting-related head impacts^21^, and we cross-referenced fight histories with online records to ensure accuracy, nof_pro may not capture unrecorded exposures such as those incurred during training. Future studies would benefit from a more comprehensive and individualized assessment of RHI exposure.

To boost the statistical power, we included both male and female retired fighters. However, the small number of female participants and sex-specific RHI responses could bias our results. Reanalysis restricted to male fighters revealed the same moderation effects of *APOE4* on longitudinal GFAP variation and *MAPT* haplotype on NfL levels (**Supplementary Table 2**). Furthermore, the *MAPT* H2 haplotype is predominantly found in Caucasians, and our inclusion of all racial groups may have introduced additional bias. Reanalyzing *MAPT*-related data in only Caucasian male fighters did not fully replicate our results, with only a trend-level significance remained for cross-sectional and longitudinal NfL variations (**Supplementary Table 2**). Future studies with larger cohorts will be necessary to further confirm these findings.

The biomarkers analyzed in this study (GFAP, NfL, and hippocampal volumes) are not specific to CTE, which limits our ability to draw direct conclusions about CTE pathology. Instead, the findings reflect genetic influences on broader RHI-related processes of neuroinflammation, axonal injury, and structural brain change. We also found no significant associations between RHI or genetic factors and plasma p-tau231 in retired fighters, suggesting that this tau marker may not capture RHI-related neurodegeneration. Given the established relevance of p-tau231 in Alzheimer’s disease (AD), these results support the view that AD and CTE involve overlapping but distinct neurodegenerative mechanisms. Future research incorporating both AD patients and healthy controls will be essential to clarify the specificity of genetic and biomarker signatures in RHI-related outcomes.

## Conclusion

In this cohort of 111 retired fighters from PABHS, *APOE4* carrier status was associated with increased vulnerability to RHI-related neuroinflammation, while the *MAPT* H2 haplotype appeared to mitigate RHI-associated axonal injury and hippocampal atrophy, suggesting partial genetic protection. These findings provide the first *in vivo* evidence that common genetic variants shape long-term trajectories of astroglial activation and neuroaxonal degeneration after repetitive head impacts, complementing post-mortem evidence implicating *MAPT* structural haplotypes in chronic traumatic encephalopathy. Incorporating genetic risk profiling into studies of RHI-related neurodegeneration may enable earlier identification of high-risk individuals and support the development of genotype-informed monitoring and prevention strategies, with replication in larger and more diverse cohorts needed to refine these applications.

### Transparency, Rigor, and Reproducibility Statement

This study is part of the PABHS. The study protocol of PABHS have been previously reported^21,22^. It included a general analytic plan, although the analysis plan for this specific article was not formally pre-registered. A total of 151 retired fighters in PABHS (as of Dec. 2024) were screened, and the final sample included 111 retired fighters with genotyping information, verified fighting histories, and at least one visit occurring two or more years post-retirement (as detailed in Fig. 1). Data for the study were collected by research staff who were blind to the relevant characteristics of the participants. The blood- and brain-based biomarker measures in the current analyses were not provided to the participants. A deidentified data set for this study is not available in a public archive. However, a deidentified participant data set and a data dictionary used to reproduce reported results can be made available upon reasonable request from a qualified investigator, subject to a signed data access agreement as allowable according to the institutional ethics board standard, by contacting the senior author (CB).

## Data Availability

All data produced in the present study are available upon reasonable request to the authors

## Authorship confirmation/contribution statement

**XZ:** conceptualization, methodology, formal analysis, visualization, writing original draft and review & editing. **DC:** methodology, visualization, and review & editing the manuscript. **LB**: data collection, visualization, and review & editing the manuscript. **ECO**: conceptualization, methodology, visualization, and review & editing the manuscript. **CB**: conceptualization, data collection, methodology, visualization, and review and editing the manuscript.

## Funding Statement

Funding for this project came from Lincy Foundation, Belator, Ultimate Fighting Championship Company (UFC), the August Rapone Family Foundation, Top Rank, Haymon Boxing, and the National Institutes of Health (NIA-grant 1RF1AG071566, NIGMS-grant P20GM109025 and NIA-grant P20AG068053).

## Author(s’) disclosure (Conflict of Interest) statement(s)

The author(s) have no competing interest to disclose.

## Ethics approval

This study is part of the PABHS that is approved by the Cleveland Clinic Institutional Review Board (IRB protocol #10-944). Written informed consent has been obtained from all participants. The protocols of the experiment have been explained to all subjects and performed according to the Declaration of Helsinki guidelines and Belmont Report.

---

**Supplementary Figure 1.** Cross-sectional associations between lifetime fight exposure (x-axis) and hippocampal volume, stratified by *APOE4* status (A, C) and *MAPT* haplotype (B, D). Left (top panels) and right (bottom panels) hippocampi are shown separately. Non-carriers (*APOE4*– or *MAPT* H1/H1) are plotted in blue; carriers (*APOE4*+ or *MAPT* H2) in red. Regression lines with 95% CI shading depict exposure–volume relationships. Violin plots display baseline hippocampal volumes by genotype. Interaction p-values are shown; FDR-significant associations are indicated with solid lines.

**Supplementary Figure 2.** Cross-sectional (A, B) and longitudinal (C, D) associations between lifetime fight exposure (x-axis) and plasma p-tau231, stratified by *APOE4* status **(A, C) and *MAPT* haplotype (B, D).** Non-carriers *(APOE4– or MAPT H1/H1)* are shown in blue; carriers (*APOE4*+ or *MAPT* H2) in red. Regression lines with 95% CI shading depict exposure-outcome relationships. Violin plots illustrate baseline distributions or annualized changes by genotype. No associations were FDR-significant.

**Supplementary Figure 3.** Schematic summary of observed associations among RHI exposure, genetic factors, and biomarker variation in retired fighters. Solid black lines indicate previously reported associations in PABHS; grey lines indicate significant main effects of RHI exposure in the current study; dashed orange lines represent moderation effects of *APOE4*; dashed purple lines represent moderation effects of *MAPT* H2. Double lines denote findings added in the present analysis. Only associations with p<0.05 are displayed; ** indicates FDR-significant associations.

**Supplementary Table 1.** Demographic and clinical characteristics of the 74 retired fighters contributing longitudinal biomarker data, stratified by *APOE4* carrier status and *MAPT* haplotype. Values are mean [SD] unless otherwise specified. Between-group differences were assessed with t-tests for continuous variables and χ² tests for categorical variables. TES = traumatic encephalopathy syndrome.

**Supplementary Table 2.** Significance of *APOE4* and *MAPT* H2 moderation effects across biomarker outcomes. P-values are shown for all retired fighters, male-only analyses, and Caucasian male-only analyses (for *MAPT*). Outcomes include baseline NfL and GFAP, annualized change (Δ) in NfL and GFAP, and annualized change in left and right hippocampal volumes. FDR-corrected results are bolded in the main text; nominal associations are reported here for completeness.

## Notes

### Competing Interest Statement

The authors have declared no competing interest.

### Author Declarations

This study is part of the PABHS approved by the Cleveland Clinic Institutional Review Board (IRB protocol #10-944).

